# The Unyvero Hospital-Acquired pneumonia panel for diagnosis of secondary bacterial pneumonia in COVID-19 patients

**DOI:** 10.1101/2020.11.24.20237263

**Authors:** Chaitanya Tellapragada, Christian G. Giske

## Abstract

The study was undertaken to evaluate the performance of Unyvero Hospitalized Pneumonia Panel (HPN) Application, a multiplex PCR based method for the detection of bacterial pathogens from lower respiratory tract (LRT) samples, obtained from COVID-19 patients with suspected secondary hospital-acquired pneumonia. Residual LRT samples obtained from critically ill COVID-19 patients with predetermined microbiological culture results were tested using the Unyvero HPN Application. Performance evaluation of the HPN Application was carried out using the standard-of-care (SoC) microbiological culture findings as the reference method. Eighty-three LRT samples were used in the evaluation. The HPN Application had a full concordance with SoC findings in 59/83 (71%) samples. The new method detected additional bacterial species in 21 (25%) and failed at detecting a bacterial species present in lower respiratory culture in 3 (3.6%) samples. Overall the sensitivity, specificity, positive and negative predictive values of the HPN Application were 95.1% (95%CI: 96.5-98.3%); 98.3% (95% CI: 97.5-98.9%); 71.6% (95% CI: 61.0-80.3%) and 99.8% (95% CI: 99.3-99.9%) respectively. In conclusion, the HPN Application demonstrated higher diagnostic yield in comparison with the culture and generated results within 5 hours.

## Introduction

Lower respiratory tract infections (LRTIs) have a diverse microbial etiology including bacterial, viral and fungal pathogens. Early and accurate identification of the infectious etiology causing LRTIs is crucial for deciding the course of treatment with reference to selection of appropriate antimicrobial therapy. Pathogen-specific antimicrobial therapy among patients with more severe form of LRTIs, such as community- and ventilator-associated pneumonia has been reported to reduce the length of hospital-stay, health care costs and adverse clinical outcomes [1]. Historically, etiological diagnosis of bacterial pneumonia has primarily been based on the microbiological culture findings. Minimum turn-around time for results from the time of sample inoculation for culture and antimicrobial susceptibility testing is approximately 36-48 hours. Sensitivity of the microbiological culture techniques for recovery of bacterial pathogens from lower respiratory tract samples can be hindered by several factors including: i) prior administration of antibiotics; ii) poor quality and low quantity of the sample; iii) overgrowth of commensal respiratory tract microbiota; iv) technical expertise of the microbiologist reading the culture plates. Semi-quantitative cultures of respiratory samples often cannot distinguish colonizer from pathogen. This problem can be overcome by using quantitative culture techniques that are more conclusive but are more laborious and cumbersome to perform.

Amidst these pre-existing challenges with reference to etiological diagnosis of LRTIs, COVID-19 has rapidly emerged as a major public health concern, worldwide. Current estimates suggest that nearly 80% of the patients admitted in the ICU with COVID-19 receive antibiotics [2]. In contrast, from a recent meta-analysis that included 24 studies from various countries, it was reported that bacterial co-infections were reported in only 7-15% of the patients admitted in the ICU [3]. Antibiotic therapy in the absence of etiological diagnosis of infection has both clinical and public health implications. Inappropriate use of antibiotics is a well-established driver for emergence of antimicrobial resistance among bacterial pathogens. Given this context, it is important to verify whether newer diagnostic modalities capable of detecting bacterial pathogens from native clinical specimens can be useful in providing early and accurate etiological diagnosis of pneumonia among critically ill COVID-19 patients [4]. Herein, we evaluated the performance of Unyvero Hospitalized Pneumonia (HPN) Application (Curetis, Germany GmbH), a multiplex PCR based method, in comparison with standard-of care microbiological culture findings for detection of bacterial pathogens from lower respiratory tract (LRT) samples obtained from critically ill COVID-19 patients.

## Material and Methods

### Study details

A laboratory-based comparative study was undertaken at the Department of Clinical Microbiology, Karolinska University Hospital, Stockholm, Sweden from April 2020 through June 2020, during the first wave of the COVID-19 pandemic in Stockholm, Sweden.

### Study samples

Lower respiratory samples frozen in the biobank of Karolinska University Laboratory were used in the present study. The biobank consisted of residual LRT samples (after standard cultures) received at the clinical microbiology laboratory during March 2020 - June 2020, obtained from hospitalized COVID-19 patients in Stockholm with a clinical suspicion of secondary bacterial infection. Microbiology culture reports, sample types and baseline demographic characteristics of the subjects were extracted from the laboratory information system using the laboratory identification numbers of the samples available in the biobank. Selection of the samples was carried out based on the following inclusion criteria: i) samples obtained from adult patients; ii) obtained from subjects admitted in the intensive-care unit; iii) samples that were either positive for normal respiratory microbiota or positive for one or more of the organism targets of the Unyvero HPN panel. Selected samples were tested with the Unyvero HPN Application.

### Standard-of-Care (SoC) testing

Quantitative culture methods were used for both invasive and non-invasive sample types at the study laboratory. Detailed description of the sample volumes, culture media used and incubation conditions for various bacterial species are described previously [5]. Results from the cultures were reported in colony-forming units (CFU) per milliliter. Identification of the bacterial species was performed using MALDI-TOF (Bruker), as per the manufacturer’s instructions. Antimicrobial susceptibility testing was performed using the disk-diffusion test and result interpretation was according to the EUCAST guidelines, version 8.0, Jan 2020 (www.eucast.org.). Detection of atypical agents such as *Mycoplasma pneumoniae, Chlamydophila pneumoniae* and *Legionella pneumophila* was performed at the study laboratory only on samples, when indicated by the clinician. For those samples, the DNA was extracted and subjected to a multiplex real-time PCR (Allplex Respiratory Panel-4, Seegene Inc.) as per the manufacturer’s instructions. Testing for *Pneumocystis jirovecii* was not indicated by the clinicians for any of the present study samples and hence not performed.

### Testing of the study samples using Unyvero HPN Application

Frozen samples were thawed for testing using the Unyvero HPN Application. Unyvero analysis was performed on the Unyvero System, consisting of a Lysator, Analyzer and Cockpit, as recommended by the manufacturer (Curetis GmbH, Holzgerlingen, Germany). The Unyvero HPN Application consisted of a sample tube with lysis buffer, a sealed master mix tube and a cartridge where the multiplex PCR is performed. List of the organisms and the antimicrobial resistance genes available on the Unyvero HPN panel for testing are shown in **Table 1**. The results were qualitatively reported as positive or negative for each organism/resistance marker.

**Table 1:**
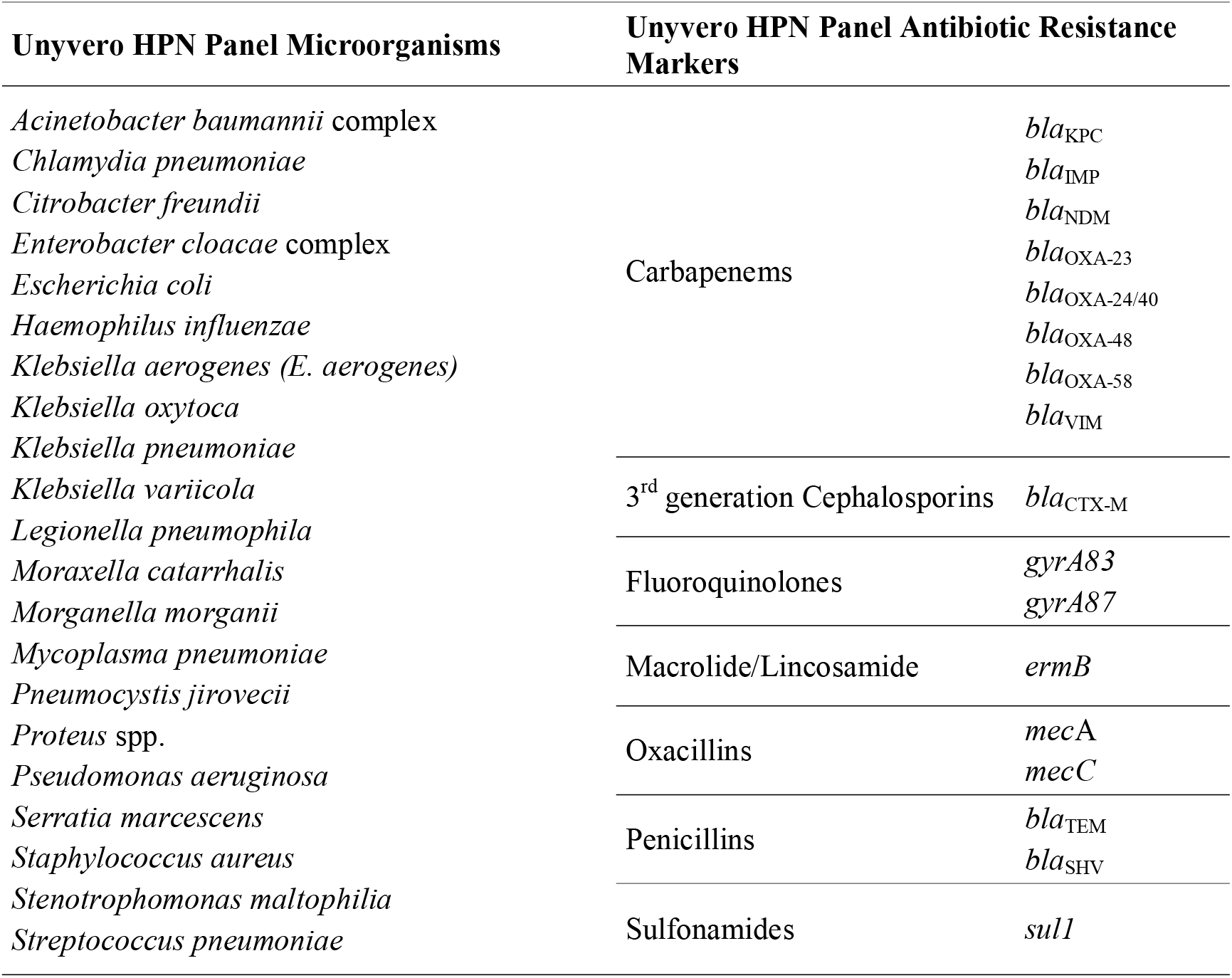
Unyvero HPN panel targets.

### Comparison of results from HPN Application results with SoC

Concordance of the HPN Application with SoC was considered when: i) same panel organism(s) detected by both HPN Application and SoC; ii) no panel organisms detected by both HPN Application and SoC; iii) no organism detected by HPN Application, culture positive for non-panel organism. Discordant results were considered when: i) same sample positive for different panel organisms by HPN Application and SoC; ii) culture positive for HPN panel organism, HPN Application negative; iii) HPN Application positive, culture-negative for the same organism. A sample was defined as both concordant and discordant if the HPN panel and SoC results had the same organism plus an additional panel organism reported by either assay with were positive according the HPN-panel cutoffs. Please refer to list of concordant and discordant samples in the supplementary section (Tables S3 – S5).

### Statistical analysis

Pathogens detected by HPN were compared to SoC results to determine overall clinical sensitivity, specificity, positive and negative predictive values (PPV and NPV, respectively) for all pathogens combined, together with confidence intervals according to the Wilson score method [6]. Sensitivity was calculated by # true positives/(# true positives + # false negatives); specificity was calculated by # true negatives /(# true negatives + # false positives); PPV was calculated by # true positives/(# true positives + # false positives); NPV was calculated by # true negatives/(# true negatives + # false negatives).

## Results

### Sample characteristics

In total, we tested 83 samples consisting of 61 (73.5%) tracheal secretions, 11 (13.4%) BAL, 8 (9.7%) PSB, 2 (2.4%) bronchial secretions and 1 (1.2%) sputum sample. The 83 samples were obtained from 68 subjects (one sample from 57 unique subjects, two samples each from 7 subjects and three samples each from 4 subjects). The multiple samples from same patient were taken on different sampling days in most cases, as indicated by the ordering clinician (**Table S1**). Seventy-four percent (50/68) of the study subjects were male and 26% (18/68) were female. Mean age of the study subjects was 58.8 ±11.3 years.

### Microbial etiology of the study samples determined using SoC

Of the total 83 samples, one bacterial species was isolated from 40 (48%) and two bacterial species were isolated from 18 (22%) samples. Normal respiratory microbiota was isolated in 25 (30%) samples. The most commonly isolated organism isolated were *S. aureus* (23/83; 27.7%) followed by *E. coli* and *K. aerogenes* in 5 (6.0%) samples each (**Table 2**). When compared against the list of organisms, available on the HPN panel, the present sample cohort comprised of 37 (44.6%) samples with one organism, 12 (14.5%) samples with two organisms and 34 (41,0%) samples with no organisms. Eleven (13.3%) samples were positive with 11 non-panel organisms (**Table S2**).

**Table 2:**
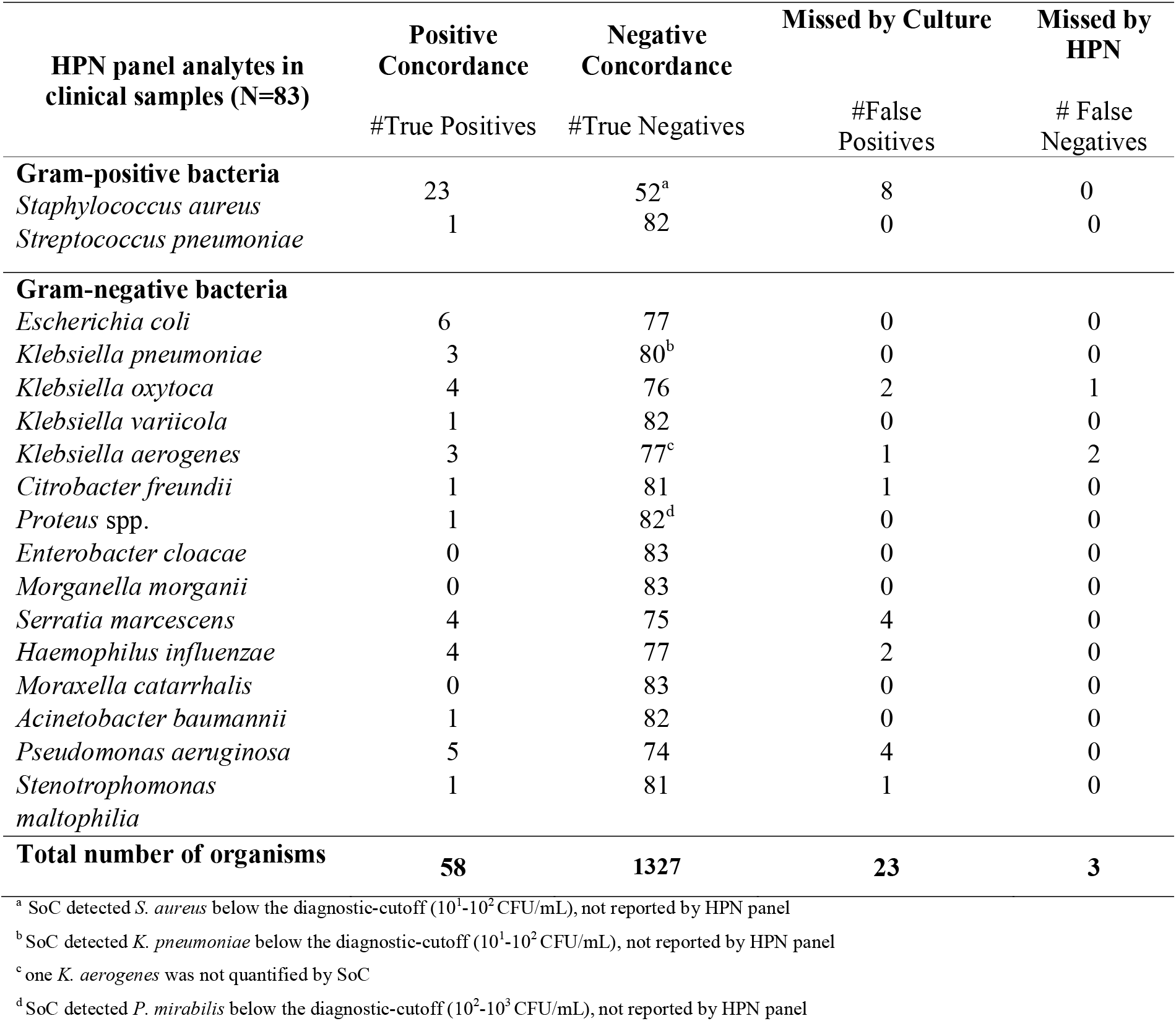
Concordance between Unyvero HPN Panel and SoC results by organism.

### Microbial etiology of the study samples determined using HPN Application

Of the 83 samples tested, one bacterial species was detected from 34 (41.4%), two bacterial species among 19 (23.1%) and three bacterial species were detected among 3 (3.6%) samples. No bacteria were detected from 27 (32.5%) samples. The most detected species were *S. aureus* (31/83; 37.4%) followed by *P. aeruginosa (*9/83; 10.9%) and *S. marcescens* (8/83; 9.6%) samples (**Table 2**).

### Comparison of results from HPN Application with SoC testing

When comparing results between HPN and SoC for individual panel analytes, sensitivity, specificity, positive predictive value and negative predictive values of the HPN application in comparison with SoC culture were: 95.1% (95%CI: 86.5-98.3%); 98.3% (95%CI: 97.5-98.9%); 71.6% (95%CI: 61.0-80.3%) and 99.8% (95%CI: 99.3-99.9%), respectively (**Table 2**). In three cases (1x *S. aureus*, 1x *Proteus* spp., 1x *K. pneumoniae*), SoC reported a pathogen concentration below the recommended reporting threshold for BAL specimens of 10^3^ CFU/mL (10). Such cases were considered “subclinical” (i.e. reporting does no merit patient treatment) and were regarded as a SoC negative result for this study. Another sample (*K. aerogenes*) which was not quantitated by SoC was therefore also regarded as SoC negative. Overall comparison of the results for concordance between both methods is shown below (**Table 3**) and sample-wise comparison of the organism yield between both methods is listed below (**Table 4**).

**Table 3:**
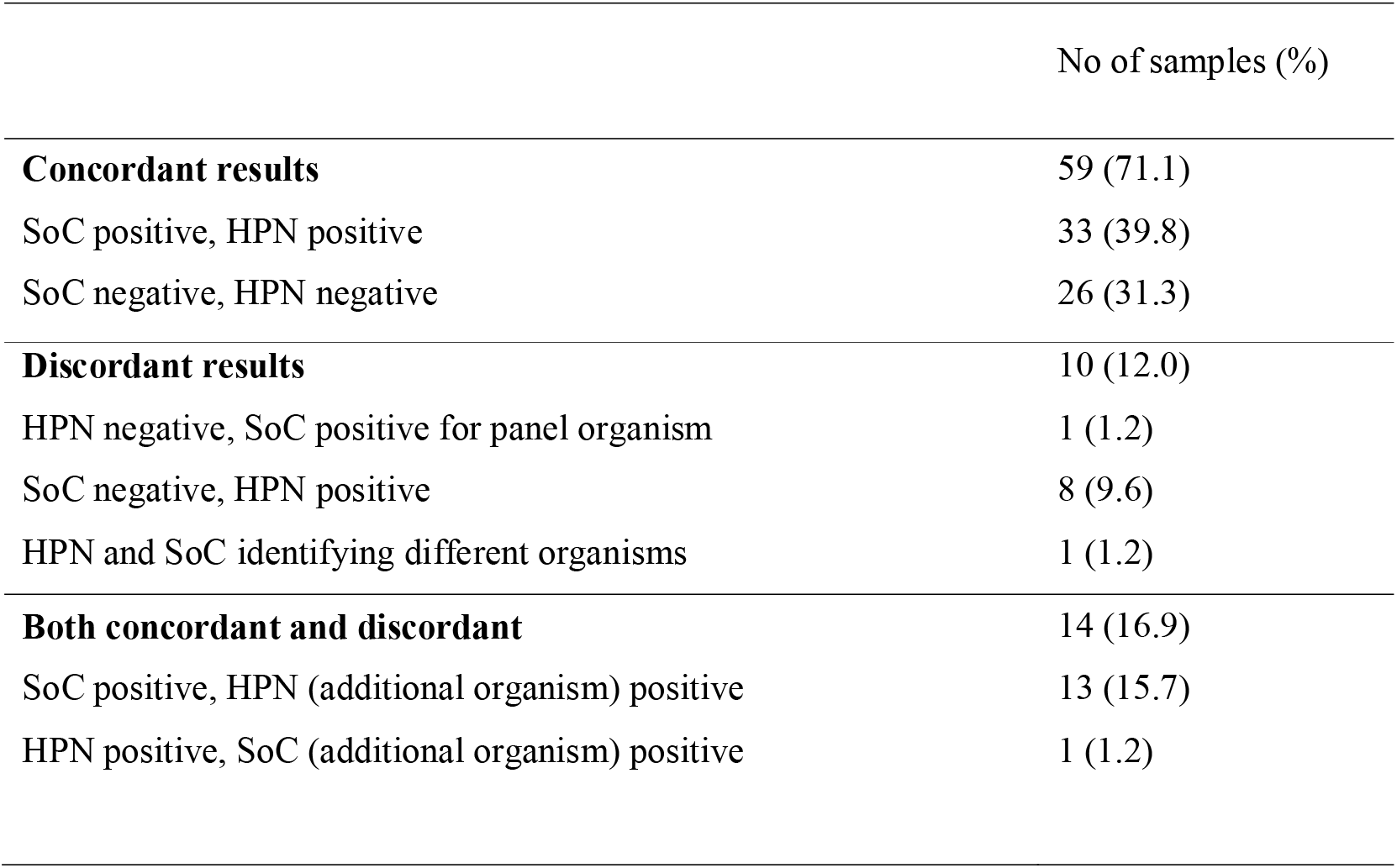
Concordance between HPN Application and SoC for HPN panel organism.

**Table 4:**
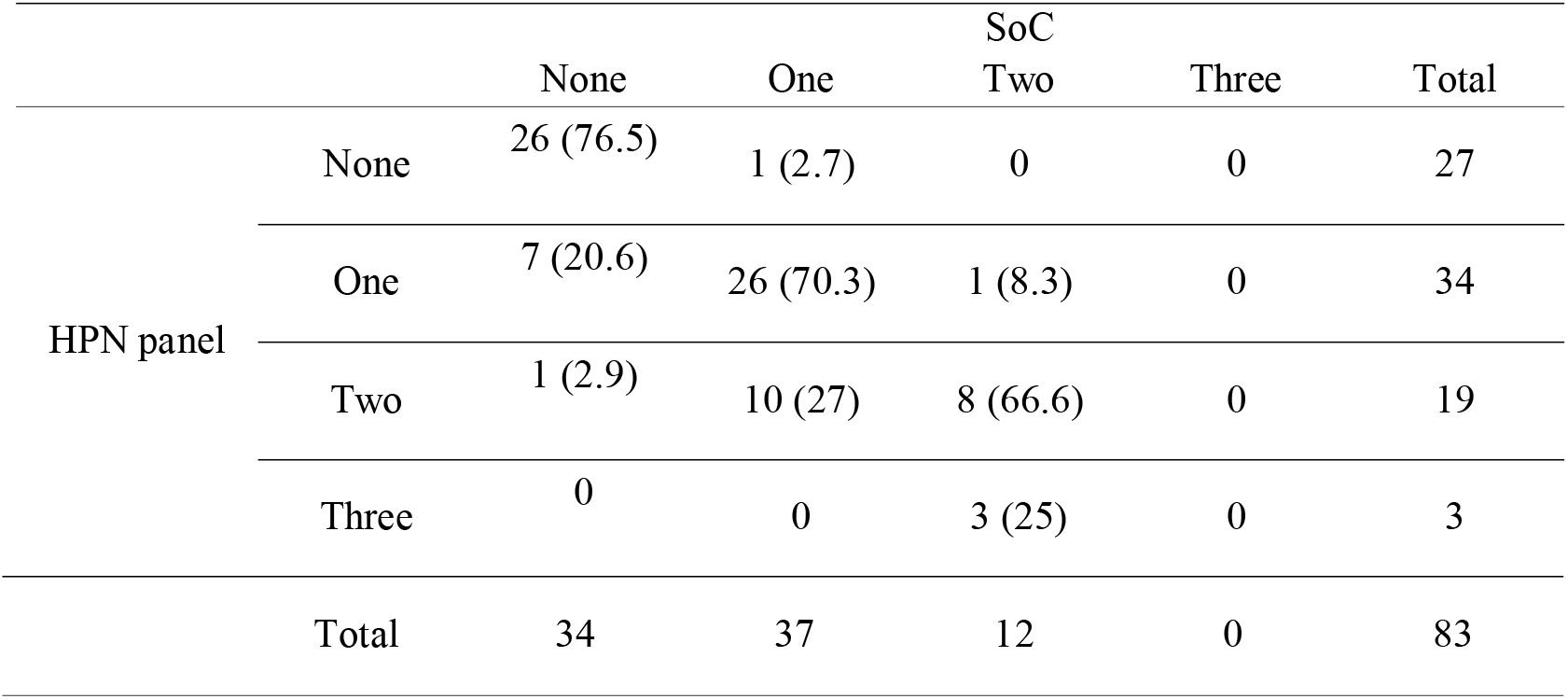
No. (%) of HPN panel organism isolated from each sample compared to SoC.

In total, the HPN Application could not detect an organism or had a discordant result in only 3/83 (3.6%) of the samples that were positive by culture for any of the panel organisms. Those three samples were: i) tracheal secretion sample that was blood-tinged, positive for *K. aerogenes* >10^5^ CFU/mL by culture and HPN did not detect any organism; ii) tracheal secretion sample positive for *K. aerogenes* >10^5^ CFU/mL by culture and HPN detected *S. aureus*; iii) BAL sample positive for *S. aureus* (10^4^ CFU/mL) and *K. oxytoca* (10^4^ CFU/mL) by culture and the HPN Application detected only *S. aureus*. On the other hand, the HPN Application concorded fully with the culture results from 59 (71.1%) samples (**Table S3**) and detected additional organisms among 21 (25.3%) samples (**Table S4**). Of the 11 non-panel organisms found on SoC (**Table S2**), 55% (6/11) were common oropharyngeal colonizing organisms that generally do not necessitate antibiotic escalation, and a further 45.5% (5/11) were organisms that would have been covered with empirical treatment.

## Discussion

The need for early initiation of pathogen-specific antimicrobial therapy among patients with severe pneumonia using rapid diagnostic techniques (RDTs) was emphasized long before the current COVID-19 pandemic began. We report here the performance characteristics of the Unyvero HPN Application for the detection of bacterial species from native lower respiratory tract (LRT) samples, with predetermined microbiological culture results, obtained from critically ill COVID-19 patients. The Unyvero HPN Application detected additional bacterial species in 25.3% (21/83) samples and failed to detect a bacterial species in only 3.6% (3/83) samples tested in the present study. Further, the new method demonstrated excellent negative predictive value (99.8%) and generated results within 5 hours from the time of loading the sample.

A recent study of the BioFire FilmArray Pneumonia panel concluded that the use of molecular diagnostic tools and the initiation of narrow-spectrum antibiotics are key elements of COVID-19 antimicrobial stewardship guidelines in critically ill [7]. Based on the recent estimates, nearly 80% of the hospitalized patients with COVID-19 are currently receiving antibiotics, albeit in the absence of a microbiological confirmation of the bacterial infection in large number of patients [2]. The rationale for antibiotic treatment in patients with COVID-19 seems to be based on the prior experience with bacterial super-infections, that were reported in nearly 11-35% patients with influenza viral infection [8]. Currently, it remains unclear whether bacterial co-infections are common among patients with SARS-CoV-2 infection, at the time of their admission to hospital; however, there is adequate evidence in the published literature suggesting that bacterial super-infections are common among COVID-19 patients admitted to the intensive-care units[2, 9, 10]. It is most likely possible that the bacterial super-infections among COVID-19 patients admitted to critical care units are due to the longer durations of stay in the ICU and mechanical ventilation, rather than the viral infection itself, but nonetheless this requires diligent microbiological testing because the signs and symptoms can be similar and confounding. Given this context, the Unyvero HPN Application can be a potential RDT of choice, considering that the HPN panel is able to detect 20 bacterial species, one fungus and 17 antimicrobial resistance genes (Table 1), that includes the most common infectious etiology of both healthcare- and ventilator-associated pneumonia.

In general, diagnostic yields from LRT samples vary with the sample type used. Invasive samples such as BAL and PSB are considered to have a better yield of the causative etiology of respiratory infections, as compared with non-invasive samples such as sputum. Diagnostic thresholds for various LRT samples among patients with health-care acquired pneumonia (HAP) range between ≥10^3^ CFU/mL for PSB samples to ≥10^5^ CFU/mL for aspirates and sputum [11]. A very low yield of pathogens from sputum samples of hospitalized patients with severe form of COVID-19 was recently reported [12]. Invasive sampling techniques have been contraindicated among COVID-19 patients outside the intensive-care units, due to the risk of aerosol generation. In the present study, tracheal aspirate was the predominant (61, 73.5%) sample type used. Nevertheless, we also included 19 (23%) BAL or PSB samples among which four (one BAL and 3 PSB) samples had a bacterial species, isolated by culture at counts ≥10^3^ CFU/mL. The Unyvero HPN Application flagged all these four samples as negative (Table 2). Considering the lower diagnostic threshold, recommended for PSB samples in comparison with other sample types (aforementioned), diagnostic efficacy of the HPN Application for PSB samples in particular, needs further evaluation.

In our study, the HPN Application detected additional bacterial species among 21/83 (25.3%) samples tested. This finding is in concordance with previous studies that have reported a similar increase in the bacterial yield from LRT samples, using other molecular detection assays among non-COVID-19 patients [13-15]. Currently, the clinical implications of detecting additional bacterial species only by the molecular methods (in the absence of culture confirmation) remain unclear and most often it is speculated that the higher yield of the molecular tests can be attributed to their ability to detect nonviable bacteria from a past infection [15]. However, in our study we also observed that the HPN Application could detect a bacterial pathogen from samples (from patients 4, 5, 9 and 11 in Table S1) that were negative by culture initially, but subsequent cultures ordered on these patients during the later course of their hospital stay were in fact positive for the same pathogen, indicating that the HPN panel can detect potential pathogens earlier than culture, which may enable earlier treatment and management of patients. Furthermore, the HPN Application demonstrated high negative predictive value of 99.8%, which would allow for reduction in unnecessary antibiotic use and support antibiotic stewardship efforts. Given this context, perhaps prospective diagnostic trials in the near future may assess the true positive predictive values of the Unyvero HPN Application and other similar commercially available molecular RDTs.

Our study has a few limitations. Currently, we do not have the clinical data of the patients from whom the present study samples were obtained. Because of this we could not determine the: i) the proportion of samples that were sent to the microbiology laboratory prior the administration of antibiotics; ii) the proportion of samples that were false positive by the HPN Application due to the detection of bacterial DNA from a past infection. Another limitation of the present study is that we could not determine the performance characteristics of the HPN Application for the detection of genes conferring antimicrobial resistance, due to the fact that only few samples yielded drug resistant phenotypes by culture in this study cohort (data not shown here). Despite these limitations, our study identified that the Unyvero HPN Application is a reliable and rapid diagnostic test with excellent negative predictive value for detection of bacterial pathogens from lower respiratory tract samples.

In conclusion, rapid diagnostics such as the Unyvero HPN panel are imperative to evaluate and test patients for bacterial pneumonia earlier in their hospital journey for more prompt and appropriate treatment. Unyvero HPN demonstrated a higher diagnostic yield than culture; it is significantly faster, with turnaround time of <5 hours from sample to results compared with average of 2.5 days for culture, providing clinicians earlier data to inform antimicrobial decisions, especially in critically ill COVID-19 patients and the upcoming flu season.

## Data Availability

All relevant data is available

## Declarations

### Ethics approval

Ethical permit was obtained from the Swedish Ethics Review Authority (Ref No:2020-04999).

### Consent to participate

Not applicable.

### Consent to publish

Not applicable.

### Data Availability

All relevant data are available in the manuscript and the supplementary files.

### Author Contributions

Both authors CT and CGG contributed to the study conception and design. The first draft of the manuscript was written by CT and CGG reviewed and approved the final version of the manuscript.

## Acknowledgements

The authors thank Curetis, Germany GmbH, for kindly providing the HPN cartridges for the present evaluation. Preliminary results were submitted as an abstract at the ESCMID Conference on Coronavirus Disease (ECCVID), September 2020.

