## Supplementary Tables for "The Unyvero Hospital-Acquired pneumonia panel for diagnosis of secondary bacterial pneumonia in COVID-19 patients"

Table S1: Results from the SoC testing and HPN Application among longitudinal samples.

| **Subject No** | **Method** | **Sample-1** | **Sample-2** | **Sample-3** | **Sampling interval between 1 and 2**  **(in days)** | **Sampling interval between 2 and 3**  **(in days)** |
| --- | --- | --- | --- | --- | --- | --- |
| **1** | SoC | *S. aureus* | *S. aureus* | NA | 20 | NA |
|  | HPN | *S. aureus* | *S. aureus* |  |  |  |
| **2** | SoC | Normal flora | *Citrobacter koseri* | NA | 7 | NA |
|  | HPN | Negative | *C. freundii* |  |  |  |
|  |  |  | *mecA* |  |  |  |
| **3** | SoC | *Citrobacter amalonaticus** | *Citrobacter amalonaticus** | NA | 0 | NA |
|  | HPN | Negative | Negative |  |  |  |
| **4** | SoC | *S. marcescens* | *S. marcescens* | NA | 6 | NA |
|  |  |  | *S. aureus* |  |  |  |
|  | HPN | *S. marcescens* | *S. marcescens* |  |  |  |
|  |  | *S. aureus* (*mec*A) | *S. aureus* |  |  |  |
| **5** | SoC | Normal microbiota | *S. aureus* | NA | 6 | NA |
|  | HPN | *S. aureus* (*mec*A, *erm*B) | *S. aureus*  (*erm*B) |  |  |  |
| **6** | SoC | *S. aureus* | Normal microbiota | NA | 4 | NA |
|  | HPN | *S. aureus* | *S. aureus* |  |  |  |
| **7** | SoC | *P. aeruginosa* | Normal microbiota | NA | 10 | NA |
|  | HPN | *P. aeruginosa* | *P. aeruginosa* |  |  |  |
| **8** | SoC | Normal microbiota | *S. aureus* | *S. aureus* |  |  |
|  | HPN | Negative | *S. aureus* | *S. aureus* | 5 | 7 |
| **9** | SoC | *Chryseobacterium indologenes** | *K. oxytoca* | Normal Flora | 7 | 5 |
|  |  |  | *C. indologenes* |  |  |  |
|  | HPN | *K. oxytoca* | *K. oxytoca* | Negative |  |  |
| **10** | SoC | *S. aureus* | Normal microbiota | Normal flora |  |  |
|  | HPN | *S. aureus* | *S. aureus* | *S. aureus* | 1 | 13 |
| **11** | SoC | Normal flora | *S. aureus* 10^3^ | Normal flora | 4 | 10 |
|  | HPN | *P. aeruginosa* | *S. aureus* | Negative |  |  |
|  |  | *S. aureus* |  |  |  |  |

*Off-panel HPN organisms

Table S2: Non-panel organisms detected by SoC. Organisms with an asterisk (*) are considered oropharyngeal flora and are not routinely treated with antibiotics when identified on culture.

| Organism | Number (%) |
| --- | --- |
| *Candida albicans** | 2 (18.2) |
| *Citrobacter amalonaticus* | 2 (18.2) |
| *Chryseobacterium indologenes* | 2 (18.2) |
| *Yeast** | 2 (18.2) |
| *Beta hemolytic Streptococcus (Group-G)** | 1 (9.1) |
| *Citrobacter koseri* | 1 (9.1) |
| *Prevotella spp** | 1 (9.1) |
| **Total non-panel organism** | **11** |

Table S3: SoC/Unyvero full concordant.

| Standard of Care culturing | HPN Application | number of specimens |
| --- | --- | --- |
| Organism | Organism |  |
| negative | negative | 26 |
| *Klebsiella aerogenes* | *Klebsiella aerogenes* | 2 |
| *Klebsiella oxytoca* | *Klebsiella oxytoca* | 3 |
| *Staphylococcus aureus* | *Staphylococcus aureus* | 10 |
| *Serratia marcescens* | *Serratia marcescens* | 2 |
| *Streptococcus pneumoniae* | *Streptococcus pneumoniae* | 1 |
| *Escherichia coli* | *Escherichia coli* | 3 |
| *Klebsiella pneumoniae* | *Klebsiella pneumoniae* | 2 |
| *Pseudomonas aeruginosa* | *Pseudomonas aeruginosa* | 2 |
| *Haemophilus influenzae* | *Haemophilus influenzae* | 2 |
| *Staphylococcus aureus* | *Staphylococcus aureus* |  |
| *Proteus* spp. | *Proteus* spp. | 1 |
| *Escherichia coli* | *Escherichia coli* |  |
| *Escherichia coli* | *Escherichia coli* | 1 |
| *Pseudomonas aeruginosa* | *Pseudomonas aeruginosa* |  |
| *Klebsiella oxytoca* | *Klebsiella oxytoca* | 1 |
| *Staphylococcus aureus* | *Staphylococcus aureus* |  |
| *Staphylococcus aureus* | *Staphylococcus aureus* | 1 |
| *Serratia marcescens* | *Serratia marcescens* |  |
| *Pseudomonas aeruginosa* | *Pseudomonas aeruginosa* | 1 |
| *Staphylococcus aureus* | *Staphylococcus aureus* |  |
| *Haemophilus influenzae* | *Haemophilus influenzae* | 1 |
| *Klebsiella pneumoniae* | *Klebsiella pneumoniae* |  |
| **total** |  | 59 |

Table S4: Unyvero additional pathogens.

| Standard of Care culturing | HPN Application | Number of specimens |
| --- | --- | --- |
| Organism | Organism |  |
| Negative | *Pseudomonas aeruginosa* | 1 |
| Negative | *Staphylococcus aureus* | 4 |
| Negative | *Klebsiella oxytoca* | 1 |
| Negative | *Pseudomonas aeruginosa* | 1 |
|  | *Staphylococcus aureus* |  |
| Negative^a^ | *Citrobacter freundii* | 1 |
| *Escherichia coli* | *Escherichia coli* | 1 |
|  | *Klebsiella aerogenes* |  |
| *Klebsiella aerogenes* | *Klebsiella aerogenes* | 1 |
|  | *Serratia marcescens* |  |
| *Stenotrophomonas maltophilia* | *Stenotrophomonas maltophilia* | 1 |
|  | *Serratia marcescens* |  |
| *Serratia marcescens* | *Serratia marcescens* | 1 |
|  | *Staphylococcus aureus* |  |
| *Pseudomonas aeruginosa* | *Pseudomonas aeruginosa* | 1 |
|  | *Staphylococcus aureus* |  |
| *Acinetobacter pittii* | *Acinetobacter baumannii* complex | 1 |
|  | *Stenotrophomonas maltophilia* |  |
| *Staphylococcus aureus* | *Staphylococcus aureus* | 1 |
|  | *Haemophilus influenzae* |  |
| *Staphylococcus aureus* | *Staphylococcus aureus* | 1 |
|  | *Pseudomonas aeruginosa* |  |
| *Staphylococcus aureus* | *Staphylococcus aureus* | 2 |
|  | *Serratia marcescens* |  |
| *Staphylococcus aureus* | *Staphylococcus aureus* | 1 |
| *Klebsiella variicola* | *Klebsiella variicola* |  |
|  | *Klebsiella oxytoca* |  |
| *Haemophilus influenzae* | *Haemophilus influenzae* | 1 |
| *Staphylococcus aureus* | *Staphylococcus aureus* |  |
|  | *Pseudomonas aeruginosa* |  |
| *Staphylococcus aureus* | *Staphylococcus aureus* | 1 |
| *Citrobacter freundii* | *Citrobacter freundii* |  |
|  | *Haemophilus influenzae* |  |
| **total** |  | 21 |

^a^SoC reported *Citrobacter koseri* (off-panel HPN organism).

Table S5: Unyvero/SoC discrepant result.

| Standard of Care culturing | HPN application | number of specimens |
| --- | --- | --- |
| Organism | Organism |  |
| *Klebsiella aerogenes* | *Staphylococcus aureus* | 1 |
| *Klebsiella aerogenes* | *negative* | 1 |
| *Staphylococcus aureus* | *Staphylococcus aureus* | 1 |
| *Klebsiella oxytoca* |  |  |
| **total** |  | 3 |
